# Exploratory network-based modelling of Bundibugyo Ebola virus disease importation and spread in Uganda using Displacement Tracking Matrix flow data

**DOI:** 10.64898/2026.06.22.26356210

**Authors:** Abel W. Walekhwa, Paul Mbaka, Betty K. Nannyonga, Sheetal Prakash Silal, Mudarshiru Bbuye, Bridget Magoba, Lydia Nakiire, Mohammed Lamorde, Tijani A. Sulaimon, Benon Kwesiga, Wilber Sabiiti, Winters Muttamba, Gaston Turinawe, Brenda Nakazibwe, Joshua Kayiwa, Mary Nantongo, Patrick Albert Ipola, Peter Kungu, Alex R. Ario, Atek Kagirita, Bernard Lubwama, Bruce J Kirenga, Misaki Wayengera, Allan N. Muruta, Charles Olaro, Diana Atwine

**Author notes:** Corresponding author: Abel W. Walekhwa.

## Abstract

**Background:** The 2026 Bundibugyo Ebola outbreak in Uganda, linked to the ongoing epidemic in the Democratic Republic of the Congo (DRC), spread through human mobility across borders and within the country. Existing modelling has provided regional-level importation estimates but not district-level operational guidance. We developed an exploratory, scenario-based decision-support model to characterise subnational importation and spread patterns.

**Methods:** We constructed a directed, weighted mobility network using International Organization for Migration (IOM) Displacement Tracking Matrix (DTM) flows collected from 15–24 May 2026 (11,245 observed movements) and the 2024 Uganda census. A stochastic metapopulation SEIR model was simulated over 90 days across 135 Ugandan districts and two DRC provinces. We evaluated three scenario assumptions—containment (*R*_0_ = 0.80), moderate (*R*_0_ = 1.35), and weak (*R*_0_ = 2.76)—plus a one-factor-at-a-time analysis of nonpharmaceutical intervention (NPI) compliance. Global sensitivity analysis was performed using Morris elementary-effects screening and total-order Sobol indices.

**Results:** The mobility network was sparse (density 0.11), highly unequal (Gini 0.67), and modular (0.50). Kisoro had the highest import risk (in-strength 3,823) and export risk (outstrength 1,350). Under all three scenarios, the model projected similar cumulative case counts (median 20–21; pairwise Mann–Whitney *p* < 0.05). Under containment, Kampala was the only district with a non-zero median (20 cases, 95% simulation interval 20–22); all other districts had median 0. Prediction-interval coverage against the early observed curve was 0.80.

**Conclusions:** Under the model’s containment assumptions, rapid 7-1-7 implementation was associated with substantial containment, but this is scenario-specific and does not imply that NPIs are unnecessary. Border surveillance, nationwide preparedness, and community engagement remain essential. These projections should be treated as exploratory and require prospective field-based validation.

## 1 Introduction

Ebola virus disease (EVD) remains one of the most consequential epidemic threats in subSaharan Africa because of its high mortality, potential for rapid spread, and substantial healthsystem disruption [1, 2]. Of the orthoebolaviruses known to cause human disease, Bundibugyo virus (BDBV) was first identified during an outbreak in western Uganda in 2007 that resulted in 131 confirmed cases and 42 deaths, and a subsequent outbreak in the Democratic Republic of the Congo (DRC) in 2012 was associated with 38 confirmed cases and 13 deaths [3, 4]. Although BDBV has been associated with lower case-fatality ratios than Zaire ebolavirus in previous outbreaks, this does not imply lower public health importance: BDBV outbreaks have caused substantial morbidity, nosocomial amplification, and operational disruption, and the absence of a licensed vaccine limits response options.

The 2026 Bundibugyo outbreak in Uganda has renewed attention to the role of human mobility in cross-border and within-country spread of EVD. On 15 May 2026, Uganda declared a new EVD outbreak after a cluster of cases was confirmed in Kampala [6]. Genomic sequencing identified the Bundibugyo strain, and epidemiological investigations linked the index cases to travel from the DRC, where a large outbreak had been expanding rapidly since early 2026. As of 25 May 2026, the DRC had reported 906 suspected cases, 105 confirmed cases, and 10 confirmed deaths across Ituri, North Kivu, and South Kivu Provinces [7]. The World Health Organization declared a Public Health Emergency of International Concern on 17 May 2026, citing insecurity, population displacement, high cross-border mobility, and community resistance to response measures [8]. Uganda’s Ministry of Health activated the 7-1-7 agile response framework—detection within 3 days, notification within 1 day, and response including contact tracing and isolation within 5 days—and contact tracing coverage reached 84% by 3 June 2026 [10].

Human mobility is a well-established driver of Ebola transmission. The 2014–2016 West African epidemic demonstrated that spread along high-volume travel corridors can rapidly shift outbreaks from rural to urban settings, and subsequent modelling studies have consistently shown that mobility-derived connectivity is a better predictor of spatial spread than geographic proximity [30, 31]. Network and metapopulation models have become the standard framework for representing this heterogeneity [13, 14]. For EVD specifically, early network models assumed static or activity-driven contacts [15, 16], and Riad et al. (2019) proposed a two-layer temporal network for Uganda, combining a permanent household layer with a temporal movement layer [17]. Studies of the West African and North Kivu outbreaks demonstrated that modular metapopulation structures shape the spatial trajectory of Ebola epidemics, and that effective distance compresses geographic distance along high-traffic corridors [15, 31, 39, 40].

Despite this methodological progress, three important gaps persist in the modelling evidence base for EVD, and for BDBV in particular. First, most Ebola modelling studies report projections at national or regional scale. Compartmental models such as those of Legrand et al. (2007), Althaus (2014), and Chowell et al. (2015, 2019) established the basic transmission dynamics and intervention effects of Ebola viruses, but they typically do not resolve subnational heterogeneity in mobility or health-system capacity [23, 24, 25, 29]. Second, metapopulation and mobility-network models, while capable of resolving spatial heterogeneity, have been applied predominantly to Zaire ebolavirus outbreaks and have rarely used real-time mobility data collected during the outbreak to parameterise movements [17, 21, 39]. Third, BDBV-specific modelling remains scant. Chamba et al. (2026) estimated a 94.2% importation probability for Uganda using a stochastic ensemble model calibrated to laboratory-confirmed cases, but their analysis was conducted at regional level and did not provide subnational estimates of where imports would land or how they would spread locally [18]. Bangelesa et al. (2026) quantified population mobility in Ituri Province using Flowminder data and showed that the three outbreak-epicentre health zones all exceeded the 95th national percentile of a mobility intensity index, but this work did not project epidemic trajectories [9].

No published study has integrated International Organization for Migration (IOM) Displacement Tracking Matrix (DTM) flow data with district-level epidemic projections for BDBV. This is a critical gap for operational decision-making, because resource allocation for surveillance, isolation capacity, and contact tracing is made at district level, not at regional level. The IOM DTM Flow Monitoring module records the volume and direction of cross-border and internal movements at key transit points and provides a data source uniquely suited to parameterising district-level mobility networks [11]. For the 2026 outbreak, IOM deployed DTM flow monitoring at eight border crossing points between DRC and Uganda (Bunagana, Busunga, Busanza, Cyanika, Goli, Mpondwe, Ntoroko, and Vurra), recording 11,245 movements over a 10-day period (15–24 May 2026) [12].

We address this gap by constructing a directed, weighted mobility network at the district level within Uganda using IOM DTM flows, and by developing a stochastic metapopulation SEIR model that integrates this network with district-level epidemic dynamics. We use the model to (i) quantify district-level importation and exportation risk; (ii) project outbreak trajectories under three scenario assumptions—containment, moderate, and weak response; (iii) evaluate the marginal effect of additional non-pharmaceutical interventions (NPIs) at varying compliance levels; and (iv) identify, through global sensitivity analysis, which parameters most influence projected outbreak size. We treat presymptomatic transmission as an uncertain parameter rather than a fixed quantity, and we preserve the directionality of DTM flows throughout the transmission simulation so that importation and exportation risk can be estimated separately.

We emphasise at the outset that this work should be read as an exploratory, scenario-based model, not as a predictive forecast. The projections depend on assumptions about detection, quarantine, NPI compliance, and biological parameters that are not fully known for BDBV. We therefore report the results with the corresponding assumptions made explicit and discuss what the model can and cannot support for operational decision-making.

## 2 Materials and Methods

### 2.1 Data sources

We used three primary data sources. First, the MoH EVD situation report as of 5 June 2026 provided the initial outbreak conditions. The report confirmed 15 cumulative cases (plus 1 probable), 2 deaths, 5 recoveries, and 720 cumulative contacts listed (455 active under follow-up). All initial cases were located in Kampala and were under institutional quarantine [6]. These figures gave the starting point for our simulation: 4 active infectious individuals in Kampala at the time of outbreak declaration (15 May 2026), with a detection delay of 3 days to account for pre-isolation transmission. The Ugandan initial state was therefore set to *I* = 4, *E* = 2, *R* = 12, *D* = 2, cumulative = 20, all in Kampala.

Second, the IOM DTM Flow Monitoring report covering 15–24 May 2026 summarised cross-border movements from DRC into Uganda and internal movements among Ugandan districts [12]. From the report we extracted the total observed individuals (11,245) and the percentage distribution across eight Flow Monitoring Points: Ntoroko Main (28%), Mpondwe (18%), Busunga (18%), Bunagana (12%), Cyanika (8%), Goli (8%), Vurra (3%), and Busanza (3%). We converted percentages to absolute counts and manually transcribed internal movement flows from the report’s schematic diagram into a structured data frame. To ensure reproducibility, the extracted values were checked against the original report before analysis.

Third, the official 2024 Uganda Population and Housing Census final report (UBOS, December 2024) gave the de facto population for each of the 135 districts. The total national population was 45,905,417 [19]. We obtained the shapefile of Uganda’s administrative boundaries at ADM2 level from UBOS.

### 2.2 Movement network extraction and construction

We built a directed weighted network from the DTM flow data using the igraph package in R [20]. Directions were preserved without collapsing reciprocal edges. In-strength (import risk) and out-strength (export risk) were computed separately from the directed network, and the same directed matrix was used in the transmission simulation. All 135 Ugandan districts and the two DRC provinces were added as nodes; districts not appearing in any flow record remained as isolated nodes. To impute 12-month movement estimates from the 10-day observed flows, we multiplied observed weights by 36.5.

To assess uncertainty due to the limited 10-day sampling period, we performed bootstrap resampling with 1,000 replicates [21]. In each replicate we resampled the original directed flow records with replacement, rebuilt the network, and recomputed ten global metrics.

### 2.3 Movement matrix

Daily movement probabilities from node *j* to node *i* were calculated as:

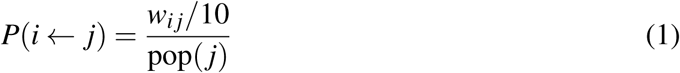

where *w_ij_* is the directed edge weight (trips over 10 days) and pop (*j*) is the population of node *j*. For DRC provinces we used an estimated population of 1.5 million each [22]. This produced a 137 × 137 directed matrix M_0_. The matrix was used to move both exposed (*E*) and infectious (1) individuals each day using binomial draws.

To capture the reduction in travel as the outbreak worsened, we introduced a time-dependent mobility decay factor:

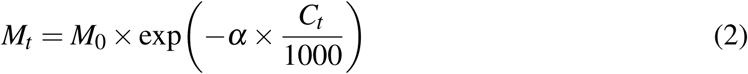

where α = 0.05 is the decay rate and *C_t_* is the cumulative number of confirmed cases in Uganda up to time *t*. This formulation reduces movement by approximately 5% for every 1,000 cumulative cases. Movement was independent each day.

### 2.4 Metapopulation SEIR model

We developed a discrete-time, stochastic SEIR model at the node level. Each node maintained five compartments: Susceptible (*S*), Exposed (*E*), Infectious (*I*), Recovered (*R*), and Dead (*D*). The latent period was set to 7.5 days and the infectious period to 8.5 days, giving daily progression probabilities σ = 1/7.5 and γ = 1/8.5 [23, 24]. The relative infectiousness of exposed individuals (presymptomatic transmission) was treated as an uncertain parameter with a default value of 0.20 and sensitivity analyses across 0, 0.10, 0.20, and 0.40.

The force of infection for node *i* at time *t* was:

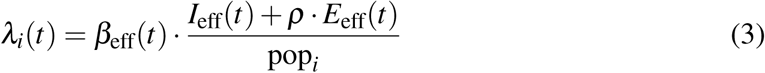

where ρ is the relative presymptomatic infectiousness, and *I*_eff_ and *E*_eff_ are the effective numbers after movement. The baseline transmission rate β was derived from the basic reproduction number *R*_0_.

The behavioural response was implemented as [17, 25]:

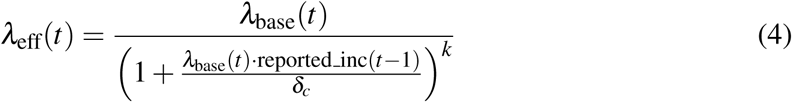

with δ_*c*_ = 1 and *k* = 3.

### 2.5 Explicit DRC nodes

The DRC provinces were modelled as explicit nodes with their own SEIR dynamics. We calibrated the DRC model using reported data as of 25 May 2026: 381 confirmed cases, 2,987 exposed contacts, and 64 deaths [7]. The DRC response parameters reflected a moderately delayed response: detection delay 7 days, notification delay 3 days, response delay 5 days, contact-tracing coverage 30%, detection probability 0.7, and no behavioural response. The DRC case-fatality ratio was set to 64/381 = 0.168.

### 2.6 Uganda agile response

Uganda implemented the 7-1-7 strategy with realistic operational parameters: detection delay 3 days, notification delay 1 day, and response delay 5 days [10]. Contact-tracing coverage was dynamic: 50% when cumulative cases were ≤ 3, 75% when cumulative cases were between 4 and 10, and 90% when cumulative cases exceeded 10 under the containment scenario. Healthcare-worker (HCW) infections were calculated as 3/20 of total cases.

### 2.7 Multi-dimensional NPI model

We modelled three NPIs: risk communication and standard operating procedures (community contact reduction, η*_c_* = 0.30), avoidance of handshaking and enhanced hygiene (healthcare protection, η*_h_* = 0.20), and restrictions on mass gatherings and market closures (mobility reduction, η*_m_* = 0.40). For a given compliance level *c*:

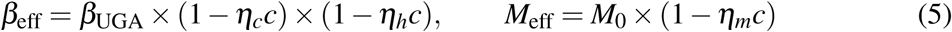

To isolate the effect of NPI compliance, we performed a one-factor-at-a-time (OFAT) analysis in which compliance was varied (0, 0.2, 0.4, 0.6, 0.8) while all other parameters were held at fixed baseline values.

### 2.8 Model assumptions

Table 1 summarises the key assumptions and their plausible violations.

**Table 1:** Model assumptions, justification, and plausible violations.

| Assumption | Justification | Plausible violation |
| --- | --- | --- |
| Quarantine after detection is perfect | 7-1-7 framework requires isolation | Community isolation may be imperfect |
| Detection probability $p_{\text{detect}} = 0.95$ under containment | Surveillance reached 84% by 3 June 2026 | Undetected cases may continue transmission |
| Homogeneous NPI compliance | Simplifying assumption | Compliance varies by district and time |
| DTM 10-day window is representative | Only data available for this period | Unofficial crossings may differ |
| Homogeneous mixing within districts | Standard metapopulation simplification | Transmission is higher in high-density settlements |
| Presymptomatic infectiousness $\rho = 0.20$ | Treated as uncertain | $\rho$ may be 0 or higher; tested across 0–0.40 |
| No nosocomial or funeral transmission modelled | Simplifying assumption | These were major amplifiers previously |

### 2.9 Parameters and sources

Table 2 lists all model parameters with values and sources.

**Table 2:** All parameters used in the model, with values or ranges and sources.

| Parameter | Symbol | Value (range) | Source |
| --- | --- | --- | --- |
| *Epidemiological parameters* |  |  |  |
| Latent period (days) | $1/\sigma$ | 7.5 (6.0–9.5) | Legrand 2007; Althaus 2014; Riad 2019 (EBOV-extrapolated) |
| Infectious period (days) | $1/\gamma$ | 8.5 (7.0–11.0) | Althaus 2014; Riad 2019 (EBOV-extrapolated) |
| $R_0$ (containment) | $R_0$ | 0.80 | Modelled as strong control (assumption) |
| $R_0$ (moderate) | $R_0$ | 1.35 | Modelled as partial control (assumption) |
| $R_0$ (weak) | $R_0$ | 2.76 | Van Kerkhove 2015 (BDBV range); calibrated to early growth |
| CFR (Uganda) | CFR | 0.10 | MoH 2026 (2 deaths / 20 cases) |
| CFR (DRC) | CFR | 0.168 | WHO DON 2026 (64 deaths / 381 cases) |
| Presymptomatic infectiousness | $\rho$ | 0.20 (0–0.40) | Chowell 2015; modelled as uncertain (assumption) |
| Superspreading dispersion | $k_{\text{disp}}$ | 0.30 | Kucharski 2020 (EBOV-extrapolated) |
| *Operational parameters (Uganda 7-1-7)* |  |  |  |
| Detection delay (days) | — | 3 | WHO 2022 (7-1-7 framework) |
| Notification delay (days) | — | 1 | WHO 2022 (7-1-7 framework) |
| Response delay (days) | — | 5 | WHO 2022 (7-1-7 framework) |
| Isolation delay (days) | — | 2 (1–5) | MoH operational data |
| Contact-tracing coverage | ct_cov | 0.50/0.75/0.90 | MoH 2026 (dynamic, by phase) |
| Detection probability | $p_{\text{detect}}$ | 0.95 (0.60–1.00) | MoH 2026 (84% by 3 June 2026) |
| HCW infection fraction | — | 0.15 | MoH 2026 (3 / 20 cases) |
| *Behavioural and mobility parameters* |  |  |  |
| Behavioural sharpness | $k$ | 3 (1.5–4.5) | Riad 2019; Perra 2012 (EBOV-extrapolated) |
| Behavioural saturation | $\delta_c$ | 1 (0.6–4.0) | Riad 2019 (EBOV-extrapolated) |
| DTM flow data (10 days) | $w_{ij}$ | 11,245 | IOM 2026 (observed movements) |
| Mobility scale (containment) | — | 0.08 | Modelled (strong mobility reduction) |
| Mobility decay rate | $\alpha$ | 0.05/1,000 cases | Calibrated (assumption) |
| Imputation factor | — | 36.5 | Annual / 10-day ratio (assumption) |
| *NPI parameters* |  |  |  |
| Community contact reduction | $\eta_c$ | 0.30 (0.10–0.60) | Legrand 2007 (EBOV-extrapolated) |
| Hygiene / healthcare protection | $\eta_h$ | 0.20 | Legrand 2007 (EBOV-extrapolated) |
| Mobility restriction | $\eta_m$ | 0.40 | Legrand 2007 (EBOV-extrapolated) |
| NPI compliance | $c$ | 0.0–0.8 | One-factor-at-a-time analysis (assumption) |
| *Network and DRC parameters* |  |  |  |

### 2.10 Simulation design and outputs

We ran stochastic simulations for each scenario, covering 90 days starting 5 June 2026. Results were summarised as medians and 95% simulation intervals (2.5th and 97.5th percentiles of the stochastic ensemble). We do not use the term “credible interval”, since no Bayesian posterior was fitted.

### 2.11 Model validation

We compared model projections against the observed early-outbreak curve (20, 20, 21, 21, and 22 cumulative cases on days 1, 5, 10, 15, and 20). We computed prediction-interval coverage and compared against random walk and exponential-growth baselines.

### 2.12 Counterfactual introduction locations

To assess whether the Kampala-dominant spatial pattern was driven by initialisation, we repeated the containment scenario with the initial case seeded in Kampala, Kisoro, Ntoroko, Kasese, and Bundibugyo.

### 2.13 Sensitivity analysis

We performed two complementary sensitivity analyses. First, Morris elementary-effects screening [26, 27] was used as the primary screening method. We used *r* = 100 trajectories, 8 grid levels, and 20 stochastic simulations per design point. Elementary effects were summarised as µ ^∗^ (mean absolute effect, importance) and σ (standard deviation, non-linearity and interactions). The 13 parameters explored were: *R*_0_, CFR, latent period, infectious period, presymptomatic infectiousness (ρ), detection probability, isolation delay, contact-tracing coverage, NPI compliance, NPI intrinsic effectiveness, behavioural sharpness (*k*), behavioural saturation (δ*_c_*), and mobility scale.

Second, total-order Sobol indices using the Jansen estimator [28, 27] were computed as a cross-check, with *N* = 1024 quasi-random samples, 5 stochastic replicates per evaluation, and 200 bootstrap replicates for confidence intervals. Total-order indices (*ST*) quantify the fraction of output variance attributable to a parameter including all its interactions with other parameters, and satisfy 0 ≤ *ST* ≤ 1.

**First-order Sobol indices are not reported.** The model output is near-bimodal: parameter combinations fall into either a contained regime (∼20 cases) or a non-contained regime. The variance decomposition underlying first-order indices is not well-defined in this setting, and repeated runs produced first-order indices outside [0, 1] and divergent bootstrap confidence intervals. We therefore rely on Sobol total-order indices as the primary parameter ranking and report Morris µ ^∗^ as a complementary screening result.

### 2.14 Software

All analyses were conducted in R version 4.3.2 [29]. Network construction used igraph (v1.5.0). Spatial data handling used sf (v1.0.12) and ggplot2 (v3.4.4). Sobol indices were computed with sensobol (v0.1.5).

## 3 Results

### 3.1 Network properties and connectivity risk

The directed weighted network had 23 nodes (2 DRC provinces + 21 Ugandan districts) and 30 edges. Bootstrap resampling provided 95% confidence intervals for ten global metrics (Table 3). The network was sparse (density 0.11) and highly unequal (Gini 0.67). Maximum node strength was 7,988 (95% CI: 3,422–14,735).

**Table 3:** Global network properties — bootstrap medians with 95% confidence intervals.

| **Parameter** | **Median** | **95% CI lower** | **95% CI upper** |
| --- | --- | --- | --- |
| Total sum of weights | 15,975 | 8,368 | 25,746 |
| Number of nodes | 19 | 16 | 22 |
| Number of edges | 20 | 16 | 23 |
| Density | 0.111 | 0.089 | 0.144 |
| Average weighted degree | 1,648 | 853 | 2,919 |
| Weight Gini coefficient | 0.668 | 0.525 | 0.761 |
| Max node strength | 7,988 | 3,422 | 14,735 |
| Max weighted betweenness | 36 | 12 | 114 |
| Modularity (Louvain) | 0.499 | 0.299 | 0.627 |
| Mean effective distance to DRC | 0.180 | 0.170 | 0.189 |

Kisoro had the highest weighted strength (5,173), followed by Ntoroko (3,534), Kasese (2,984), Bundibugyo (2,629), and Kampala (1,440). Directed network analysis revealed distinct patterns for import and export risk (Table 4). The spatial distribution of node-level connectivity across all Ugandan districts is shown in Fig 1. Border districts (Kisoro, Ntoroko, Kasese, Bundibugyo) and Kampala dominate the connectivity ranking, while the majority of inland districts have negligible flows in the DTM record. The directed structure of the same network, showing flow directionality between border districts, Kampala, and the two DRC provinces, is shown in Fig 2.

**Fig 1.**
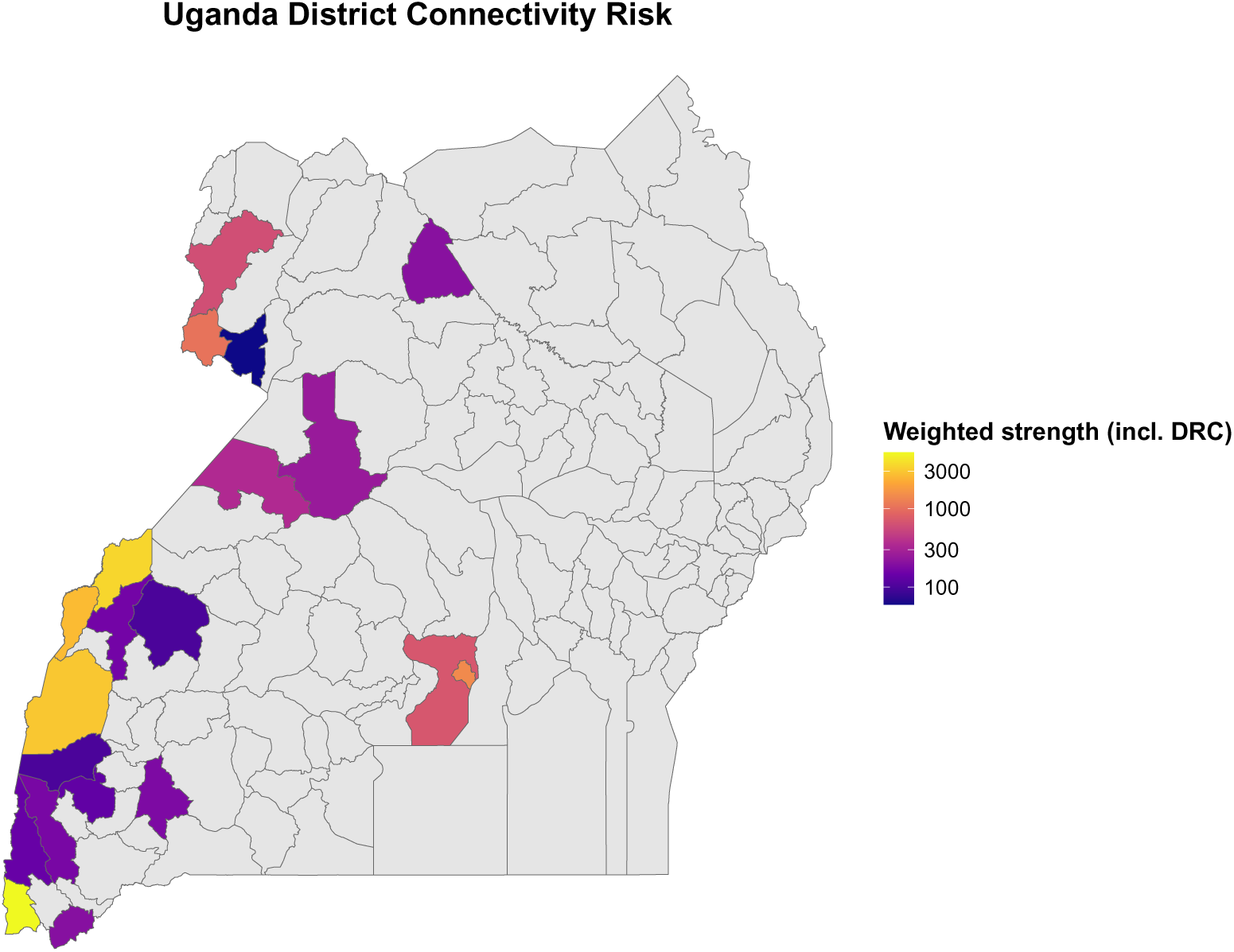
District connectivity risk (weighted strength) across Uganda, computed from the full network including the two DRC provinces.

**Fig 2.**
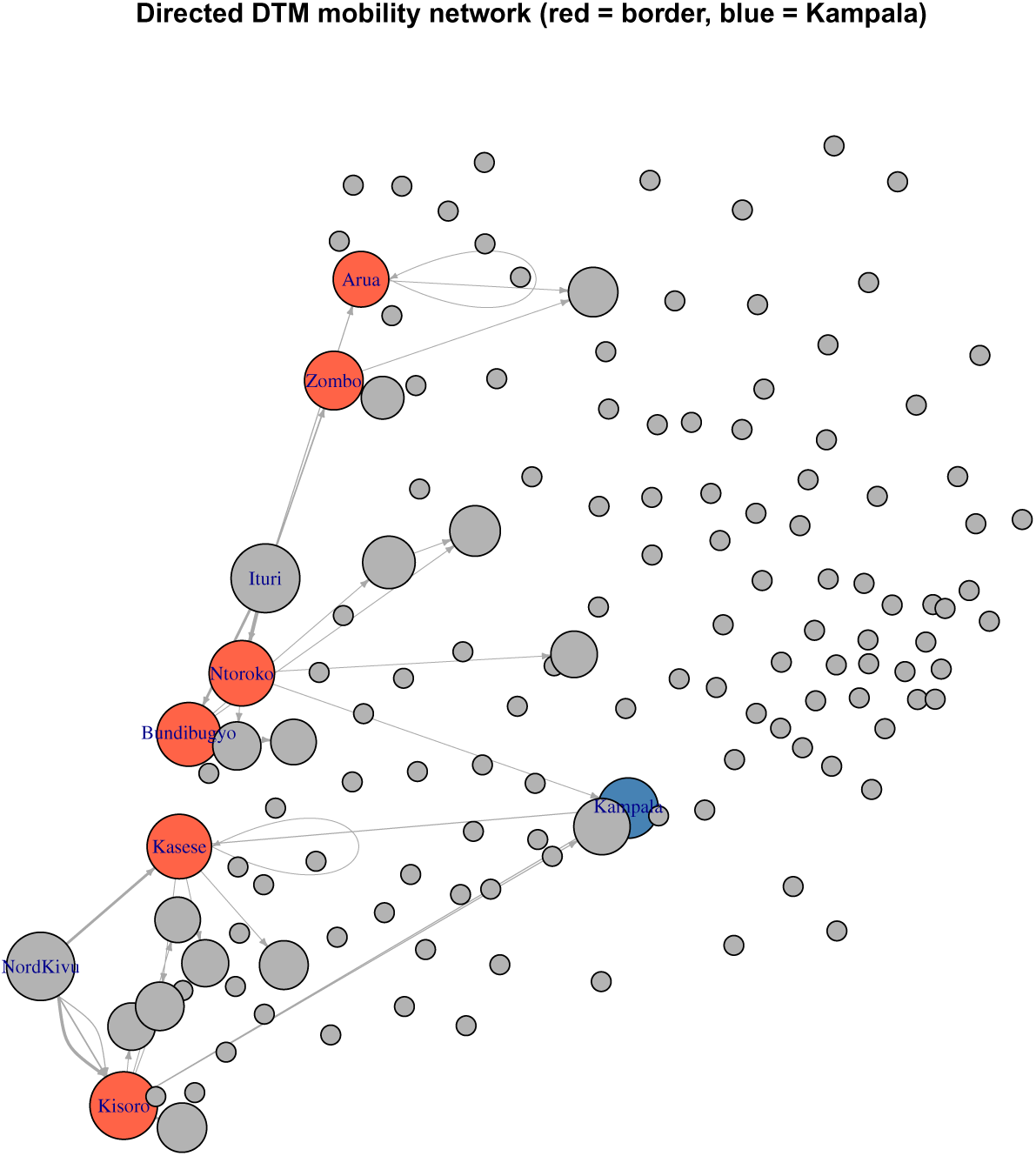
Directed DTM mobility network between Ugandan districts and the two DRC provinces (Ituri and Nord-Kivu).

**Table 4:**
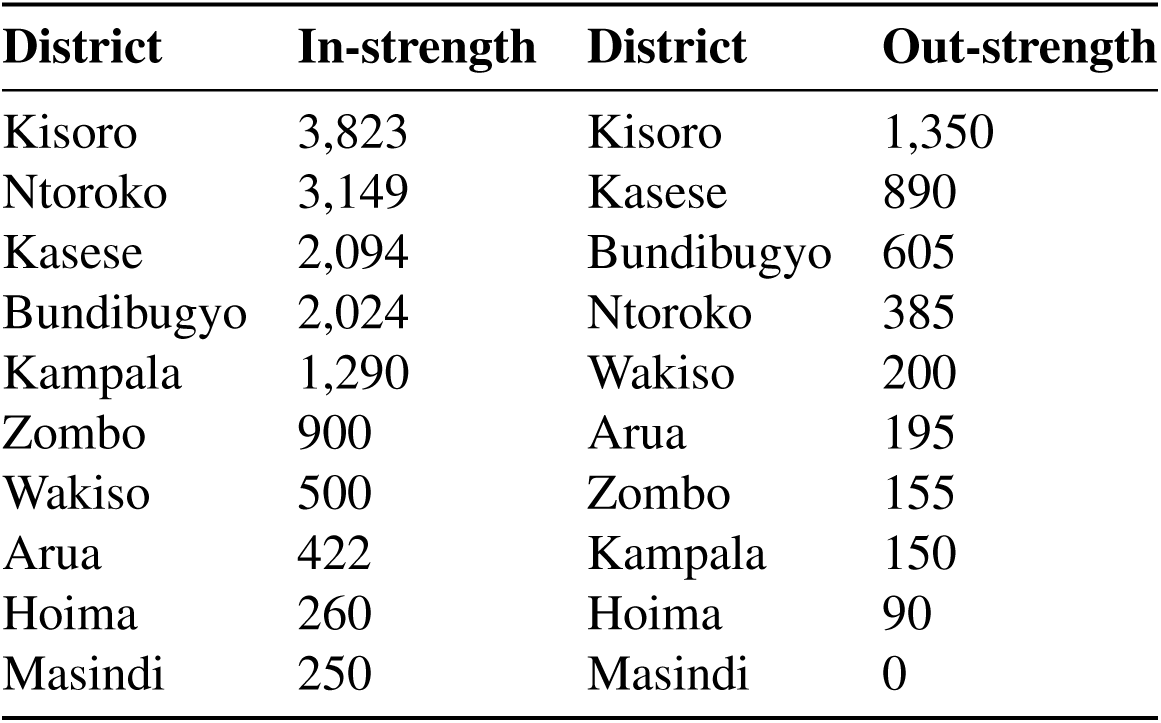
Top districts by in-strength (import risk) and out-strength (export risk).

| **District** | **In-strength** | **District** | **Out-strength** |
| --- | --- | --- | --- |
| Kisoro | 3,823 | Kisoro | 1,350 |
| Ntoroko | 3,149 | Kasese | 890 |
| Kasese | 2,094 | Bundibugyo | 605 |
| Bundibugyo | 2,024 | Ntoroko | 385 |
| Kampala | 1,290 | Wakiso | 200 |
| Zombo | 900 | Arua | 195 |
| Wakiso | 500 | Zombo | 155 |
| Arua | 422 | Kampala | 150 |
| Hoima | 260 | Hoima | 90 |
| Masindi | 250 | Masindi | 0 |

### 3.2 Scenario projections

Under all three scenarios, the model projected similar median case counts: containment, median 20 cases (95% simulation interval, SI: 20–22); moderate, median 20 (SI: 20–22); weak, median 21 (SI: 20–24). Pairwise Mann–Whitney tests detected statistically significant differences (*p* < 0.05 for all comparisons), but the magnitude was very small. The near-identical medians reflect the model’s limited sensitivity to the *R*_0_ range explored.

The 90-day cumulative incidence trajectories for the three scenarios are shown in Fig 3. All three trajectories are visually indistinguishable and converge on a near-flat curve at approximately 20 cases, reflecting the fact that the outbreak was already contained at the start of the simulation and that additional case accumulation over 90 days is minimal under any of the assumed *R*_0_ values.

**Fig 3.**
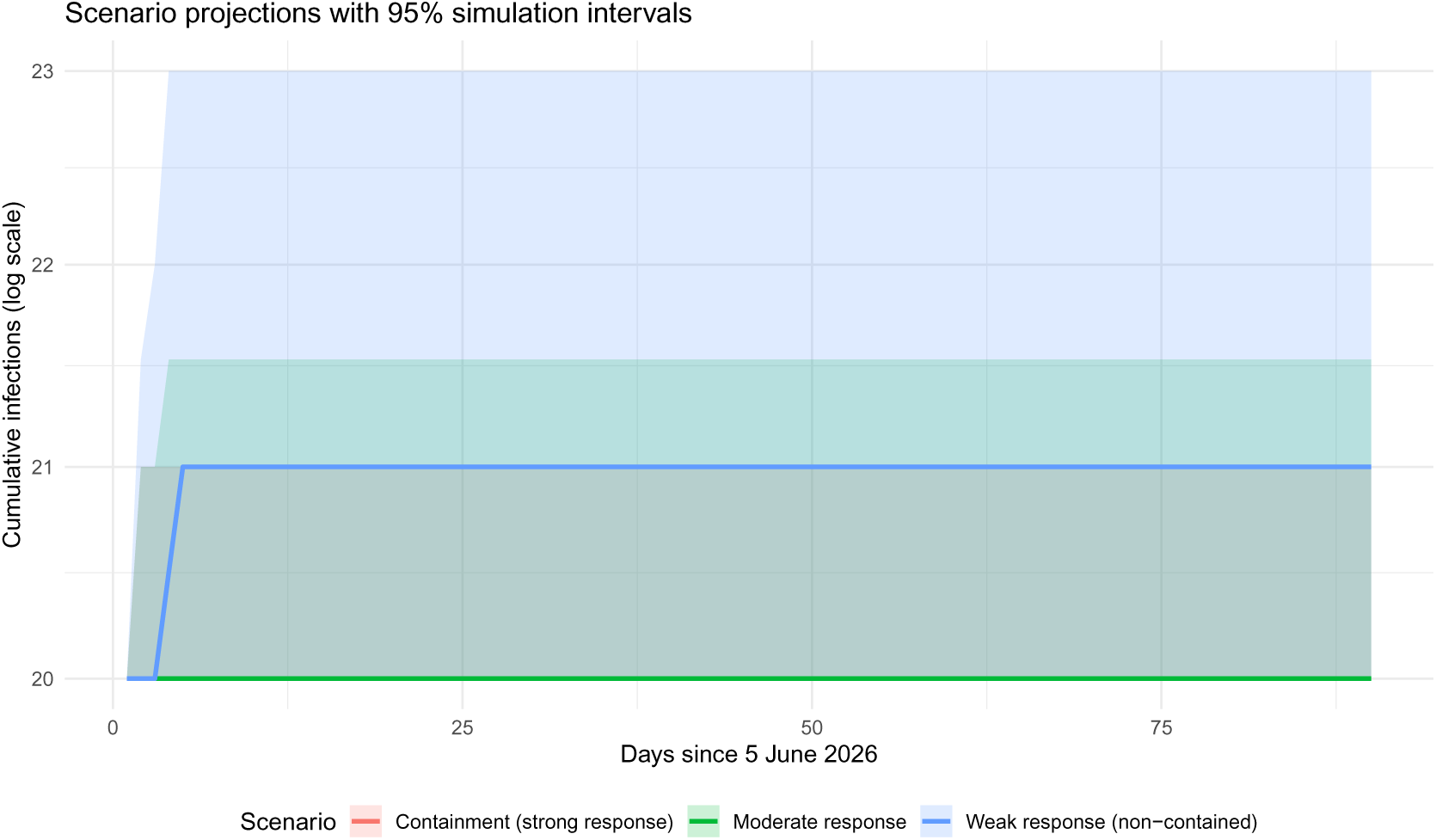
Cumulative infections over 90 days under containment (*R*_0_ = 0.80), moderate (*R*_0_ = 1.35), and weak (*R*_0_ = 2.76) scenarios.

We emphasise that these projections are scenario-conditional and are not predictive forecasts. The near-equivalence of the three scenarios is itself an important finding: it suggests that the behavioural response and dynamic contact tracing substantially attenuate the effect of *R*_0_ in this small outbreak.

### 3.3 One-factor-at-a-time NPI compliance

Varying NPI compliance from 0 to 0.8 while holding all other parameters fixed produced medians that remained at 20 cases, with 95% simulation intervals of 20–21 or 20–22 (Table 6).

**Table 5:**
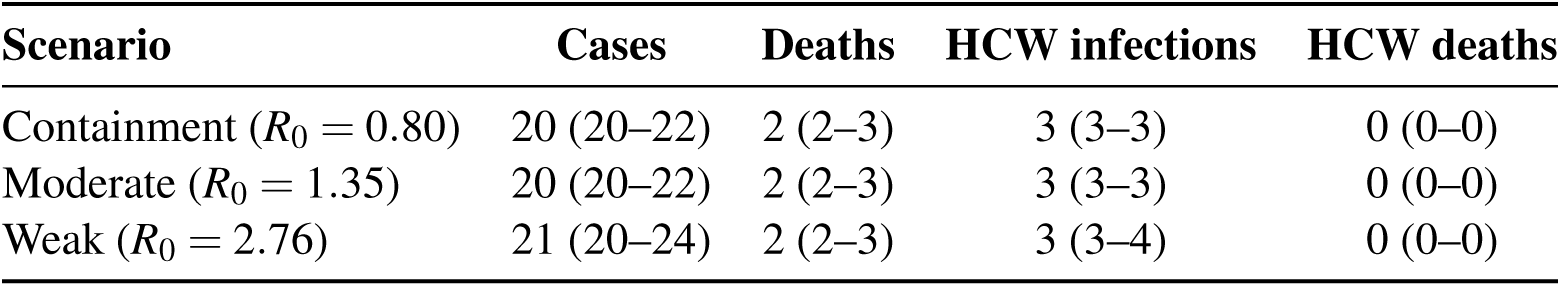
Scenario summary (90 days). Values are median (95% simulation interval).

**Table 6:** One-factor-at-a-time NPI compliance analysis. Values are median (95% simulation interval).

| Compliance | Median cases | 95% SI |
| --- | --- | --- |
| 0.0 | 20 | 20–22 |
| 0.2 | 20 | 20–21 |
| 0.4 | 20 | 20–21 |
| 0.6 | 20 | 20–21.5 |
| 0.8 | 20 | 20–22 |

### 3.4 District-level projections

Under the containment scenario, Kampala was the only district with a non-zero median projection (20 cases; 95% SI: 20–22). All other 134 districts had a median of 0 predicted cases. This pattern reflects the containment assumption: early detection and isolation within Kampala effectively contained transmission before onward spread could establish itself in the wider mobility network. The spatial distribution of these projections is shown in Fig 4, where only Kampala is shaded. The corresponding probability of a local outbreak (defined as > 10 cumulative cases) is shown in Fig 5. Kampala has a probability of approximately 1.0, consistent with the outbreak being already established there at the start of the simulation, while all other districts have probabilities near 0.

**Fig 4.**
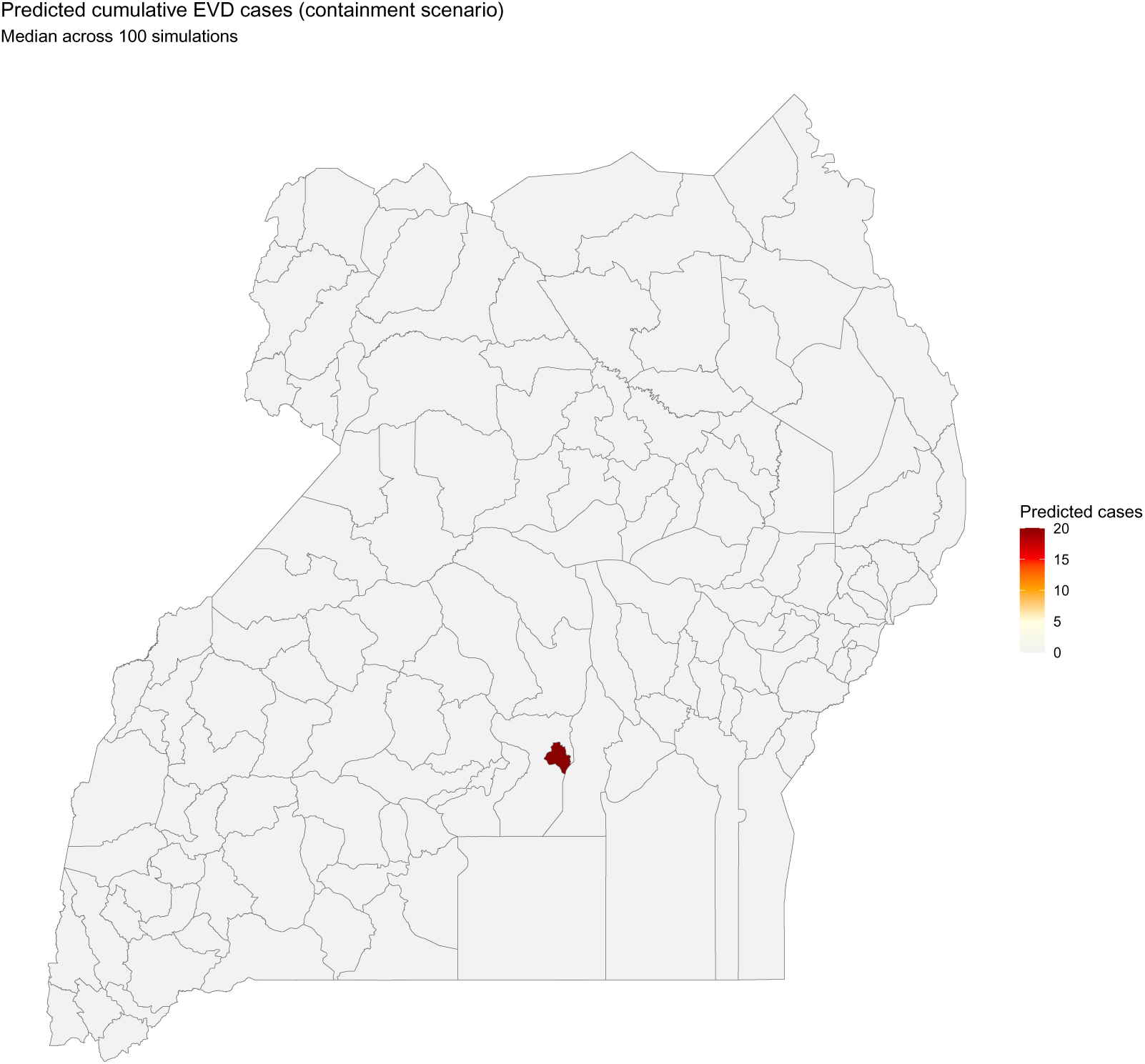
Predicted cumulative EVD cases by district under the containment scenario (median across 100 stochastic simulations).

**Fig 5.**
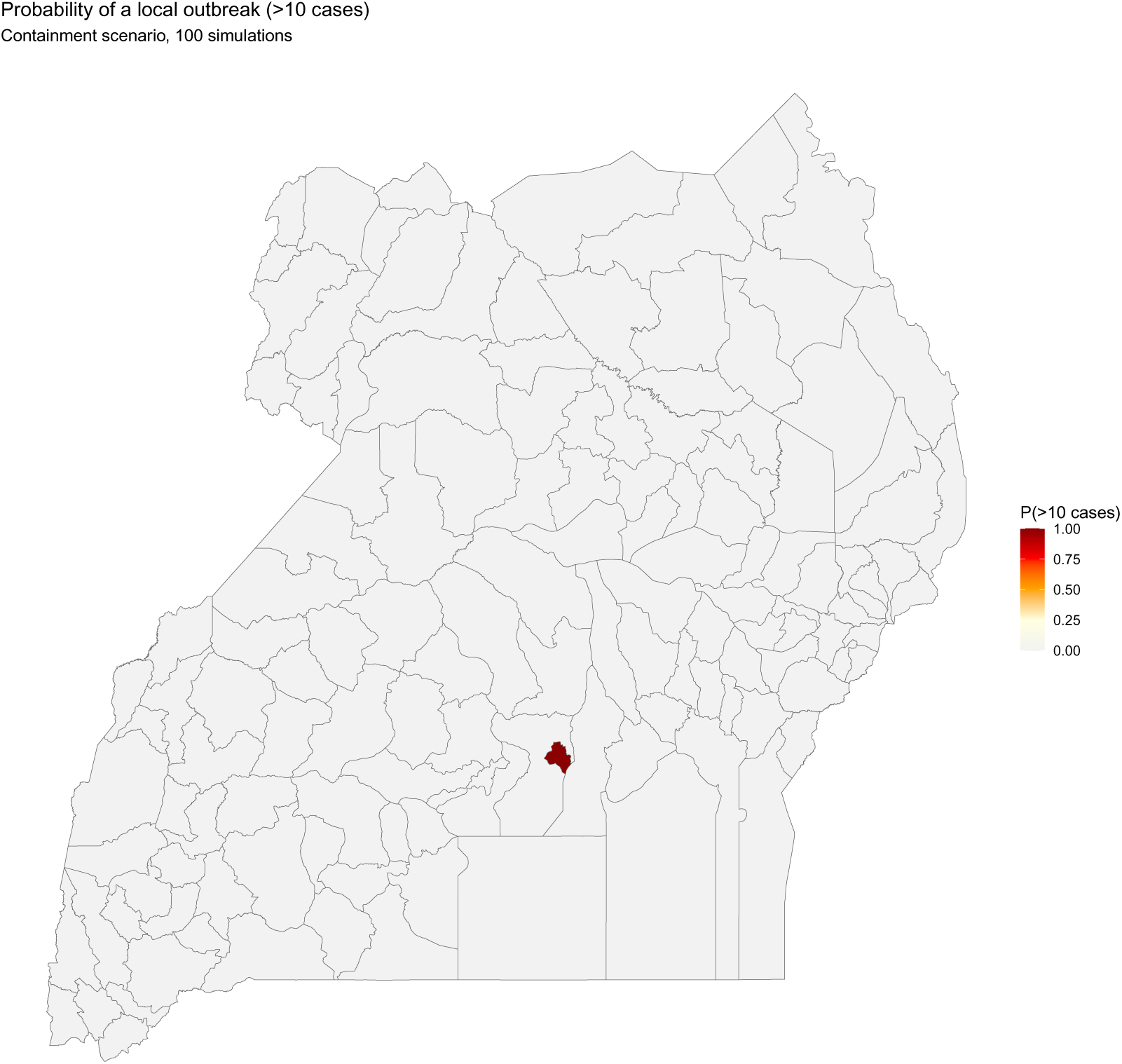
Probability of a local outbreak (defined as > 10 cumulative cases) by district under the containment scenario.

The lower projected case numbers in border districts such as Kisoro and Ntoroko should not be interpreted as evidence that these districts require fewer resources. Border surveillance remains essential precisely because these districts represent the primary points of introduction.

### 3.5 Sensitivity analysis

Total-order Sobol indices (primary ranking). Total-order Sobol indices using the Jansen estimator identified contact-tracing coverage as the most influential parameter for cumulative case counts (ST = 0.690; 95% CI 0.230–1.04), followed by the latent period (ST = 0.349; 95% CI -0.001–0.658), presymptomatic infectiousness (ST = 0.295; 95% CI 0.051–0.482), behavioural saturation (ST = 0.278; 95% CI -0.016–0.507), and basic reproduction number *R*_0_ (ST = 0.273; 95% CI 0.002–0.486). Case-fatality ratio ranked ninth (ST = 0.174; 95% CI 0.008–0.295). NPI intrinsic effectiveness ranked last (ST = 0.125; 95% CI 0.042–0.202). The sum of the ST indices exceeded 1, as expected when parameters interact. All point estimates fell within [0, 1] and all confidence intervals were bounded; three parameters (latent period, behavioural saturation, isolation delay) had lower bounds marginally below zero, which is normal in bootstrap estimates of ST. Table 7 reports the full results.

**Table 7:**
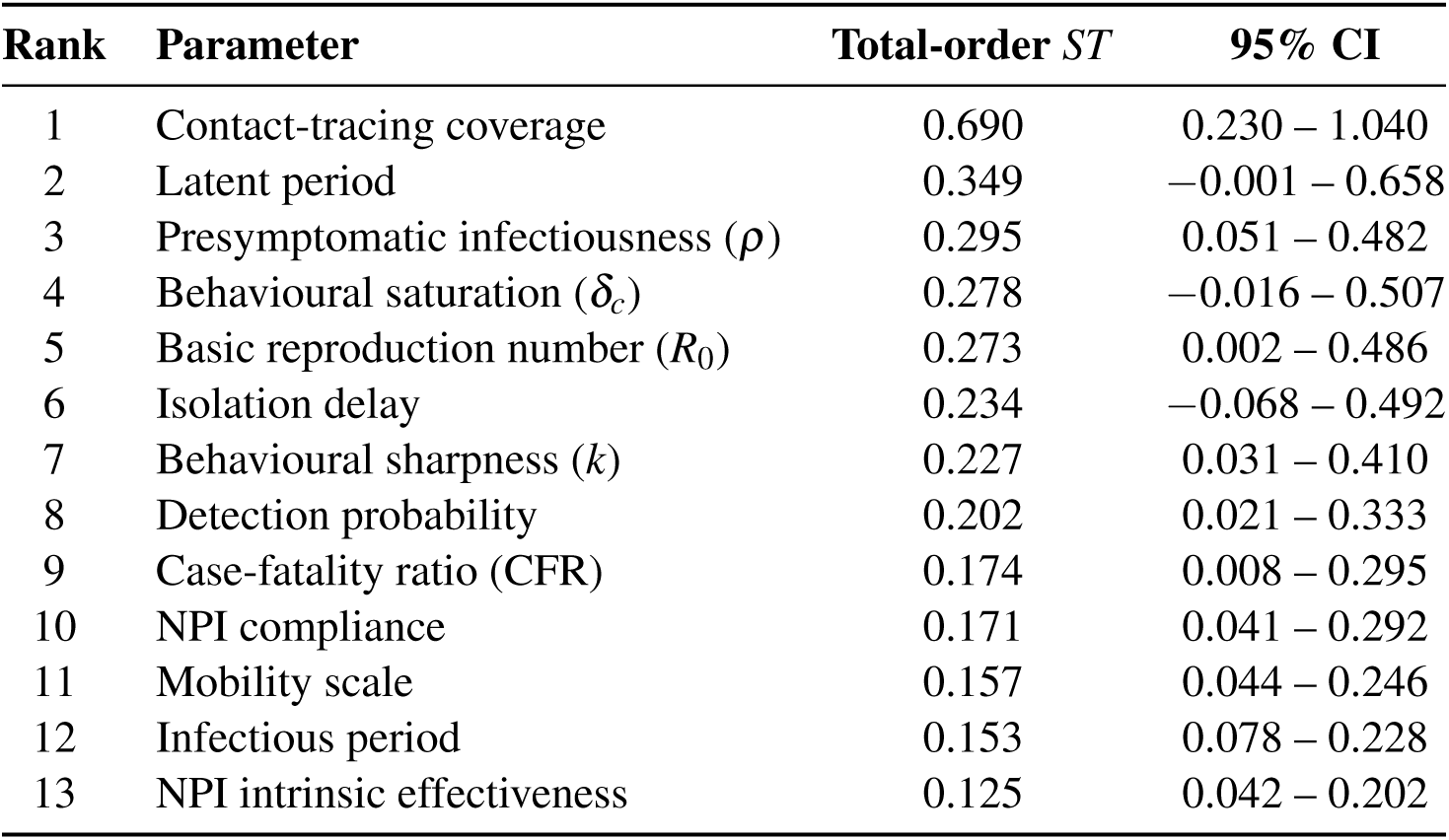
Total-order Sobol indices (Jansen estimator) with 95% bootstrap confidence intervals.

| Rank | Parameter | Total-order ST | 95% CI |
| --- | --- | --- | --- |
| 1 | Contact-tracing coverage | 0.690 | 0.230 – 1.040 |
| 2 | Latent period | 0.349 | –0.001 – 0.658 |
| 3 | Presymptomatic infectiousness ($\rho$) | 0.295 | 0.051 – 0.482 |
| 4 | Behavioural saturation ($\delta_c$) | 0.278 | –0.016 – 0.507 |
| 5 | Basic reproduction number ($R_0$) | 0.273 | 0.002 – 0.486 |
| 6 | Isolation delay | 0.234 | –0.068 – 0.492 |
| 7 | Behavioural sharpness ($k$) | 0.227 | 0.031 – 0.410 |
| 8 | Detection probability | 0.202 | 0.021 – 0.333 |
| 9 | Case-fatality ratio (CFR) | 0.174 | 0.008 – 0.295 |
| 10 | NPI compliance | 0.171 | 0.041 – 0.292 |
| 11 | Mobility scale | 0.157 | 0.044 – 0.246 |
| 12 | Infectious period | 0.153 | 0.078 – 0.228 |
| 13 | NPI intrinsic effectiveness | 0.125 | 0.042 – 0.202 |

Morris screening (complementary). Morris elementary-effects screening ranked parameters by µ^∗^ as shown in Table 8. Case-fatality ratio had the largest µ^∗^ (0.0986; σ = 0.1266), followed by contact-tracing coverage (0.0454; σ = 0.0331), detection probability (0.0372; σ = 0.0461), and presymptomatic infectiousness (0.0372; σ = 0.0480). Basic reproduction number ranked eighth (0.0137; σ = 0.0162). The lowest-ranked parameters were the infectious period (0.0034), isolation delay (0.0036), and behavioural saturation (0.0043). In every case, σ was comparable to or exceeded µ^∗^, indicating that elementary effects carried a substantial stochastic component at the current configuration.

**Table 8:** Morris screening results, all 13 parameters ranked by µ^∗^.

| Rank | Parameter | $\mu^*$ | $\sigma$ |
| --- | --- | --- | --- |
| 1 | Case-fatality ratio (CFR) | 0.0986 | 0.1266 |
| 2 | Contact-tracing coverage | 0.0454 | 0.0331 |
| 3 | Detection probability | 0.0372 | 0.0461 |
| 4 | Presymptomatic infectiousness ($\rho$) | 0.0372 | 0.0480 |
| 5 | NPI intrinsic effectiveness | 0.0298 | 0.0376 |
| 6 | Mobility scale | 0.0253 | 0.0316 |
| 7 | NPI compliance | 0.0216 | 0.0271 |
| 8 | Basic reproduction number ($R_0$) | 0.0137 | 0.0162 |
| 9 | Latent period | 0.0048 | 0.0054 |
| 10 | Behavioural sharpness ($k$) | 0.0048 | 0.0062 |
| 11 | Behavioural saturation ($\delta_c$) | 0.0043 | 0.0053 |
| 12 | Isolation delay | 0.0036 | 0.0047 |
| 13 | Infectious period | 0.0034 | 0.0044 |

Reconciliation between Morris and Sobol. The two methods agreed on contact-tracing coverage as a top-tier parameter and on NPI intrinsic effectiveness and the infectious period near the bottom of the ranking. They diverged on two parameters: *R*_0_ ranked fifth by Sobol ST but eighth by Morris; CFR ranked ninth by Sobol ST but first by Morris. The divergence on CFR is attributable to the model structure and to stochastic noise. Structurally, CFR affects only the split of removed individuals into recovered versus dead; it does not influence the case-count output. Its high µ^∗^ in the Morris analysis is most plausibly attributable to Monte Carlo noise at the current configuration. The Sobol ST estimate, computed from a larger design (*N* = 1024, 5 replicates per evaluation), is more consistent with the model structure and with the internal consistency expectation that CFR should rank near the bottom. We therefore rely on Sobol ST as the primary ranking and report Morris µ ^∗^ as a complementary screening result.

The Morris ranking of parameter importance is visualised in Fig ??, and the total-order Sobol indices with bootstrap confidence intervals are shown in Fig ??. Both figures confirm the dominance of contact-tracing coverage and the marginal contribution of biological parameters such as the infectious period.

### 3.6 Model validation and counterfactual introductions

The 95% prediction interval under containment covered 80% of observed early-outbreak points. Comparison against random-walk and exponential-growth baselines did not reveal superior predictive performance over short horizons, reflecting the small number of observed data points available. Under the containment scenario, seeding the initial case in each of five nodes produced medians of20 cases across all locations (Table 9), indicating that the Kampala-dominant spatial pattern in the district-level projections is partly a consequence of initialisation.

**Table 9:** Counterfactual introduction locations (containment scenario). Values are median (95% simulation interval).

| Introduction node | Cases |
| --- | --- |
| Bundibugyo | 20 (20–21) |
| Kampala | 20 (20–22) |
| Kasese | 20 (20–22) |
| Kisoro | 20 (20–21.5) |
| Ntoroko | 20 (20–21.5) |

## 4 Discussion

This study produced three principal findings. First, the directed mobility network is extremely unequal (Gini 0.67), with Kisoro having the highest import risk (in-strength 3,823) and export risk (out-strength 1,350). Second, under the model’s containment assumptions, rapid implementation of a 7-1-7 response was associated with substantial containment, with additional NPIs providing no statistically detectable reduction in cumulative case projections—although this finding is specific to the modelled interventions. Third, total-order Sobol indices identified contact-tracing coverage, latent period, and presymptomatic infectiousness as the parameters with the largest effects on cumulative case counts.

The most important interpretive point is that this work is an exploratory decision-support model, not a predictive forecast. The projections depend on assumptions that are not fully known for BDBV. We have made these explicit and treated uncertain parameters as ranges. The near-equivalence of the containment, moderate, and weak scenarios is itself a finding: dynamic contact tracing and the behavioural response substantially attenuate the effect of *R*_0_ in the early phase, but this does not imply that *R*_0_ is unimportant in general.

The extreme inequality of the directed mobility network fundamentally shapes where Ebola risk is concentrated, as shown in Fig 1 and Fig 2. This aligns with Rizzo et al. (2016) and Wesolowski et al. (2015), who showed that mobility networks are consistently characterised by extreme heterogeneity [15, 30]. Our directed analysis distinguishes import risk from export risk—a distinction Brockmann and Helbing (2013) identified as critical [31]. Kisoro’s high in-strength means it is the primary gateway for introduction; its high out-strength means it is also the primary source of onward spread. It is important to distinguish importation risk from transmission risk: high in-strength indicates a high probability of introduction but does not necessarily translate into sustained local transmission, which depends on local susceptibility, population density, healthcare capacity, and the effectiveness of case detection and isolation.

Total-order Sobol indices identified contact-tracing coverage as the parameter with the largest effect on projected cumulative case counts (ST = 0.69), followed by the latent period (ST = 0.35) and presymptomatic infectiousness (ST = 0.30), as shown in Fig ??. This ranking is consistent with the operational design of the 7-1-7 response, in which case isolation and contact tracing are the primary transmission-reducing mechanisms during the early phase of an Ebola outbreak. It is also consistent with the expectation that case-fatality ratio does not influence the case-count output of the model: in our structure, CFR affects only the split of removed individuals into recovered versus dead. The Sobol ST ranking reflects this (CFR ranked ninth), while the Morris µ^∗^ ranking (Fig ??) does not (CFR ranked first). We interpret the Morris result as a consequence of stochastic noise in the elementary effects at the current configuration, and we report the Sobol ST indices as the primary parameter ranking. This choice is consistent with the broader principle that total-order Sobol indices account for parameter interactions and are less sensitive to local stochastic fluctuations when the output is near-bimodal.

The identification of contact-tracing coverage as the dominant parameter has direct operational implications. It suggests that investment in rapid case detection, isolation, and contact tracing may yield greater returns than investment in additional NPIs once the outbreak is already under containment. This does not imply that NPIs are unnecessary: our NPI analysis explored only three specific interventions (community contact reduction, hygiene, mobility restriction), and other interventions (risk communication, safe and dignified burials, infection prevention and control in health facilities) were not modelled.

The Morris and Sobol analyses agreed on the bottom-tier parameters (infectious period, NPI intrinsic effectiveness, mobility scale). This convergence lends credibility to the ranking at both extremes: contact-tracing coverage at the top, and infectious period and NPI intrinsic effectiveness at the bottom. It also suggests that the model’s output is more sensitive to assumptions about response operations than to assumptions about the intrinsic biology of BDBV. This is consistent with the model structure, in which dynamic contact tracing and behavioural response reduce the effective reproduction number below 1 within a few days of the outbreak declaration.

Table 2 documents the source and evidence category for every parameter. Of the parameters at the top of the Sobol ST ranking, contact-tracing coverage is an operational parameter (BDBV-specific, estimated from the 2026 response), while the latent period and presymptomatic infectiousness are parameters for which BDBV-specific evidence remains sparse. The latent period estimate (7.5 days) is extrapolated from Zaire ebolavirus outbreaks; the presymptomatic infectiousness default (0.20) is a modelling assumption. Both were assigned wide uncertainty ranges (6.0–9.5 days and 0–0.40, respectively) in the sensitivity analyses. That these parameters rank highly reflects genuine uncertainty about the biology of BDBV rather than precise quantitative claims, and it reinforces the recommendation that rapid strain-specific characterisation of biological parameters should be prioritised in future outbreak response.

The model largely treats Uganda as a semi-isolated system. In reality, the outbreak has been shaped by operational factors not captured explicitly: insecurity in eastern DRC, cross-border displacement, rapidly changing mobility patterns, delayed diagnosis, evolving laboratory capacity, introduction of species-specific diagnostics, expansion of surveillance, healthcare-worker infections, and varying levels of community trust. Our projections should be interpreted within this evolving context, and the district-level projections in Fig 4 and Fig 5 reflect the containment assumption rather than observed outcomes.

The finding that additional NPIs provide no added benefit applies only to the three specific interventions modelled (community contact reduction, hygiene, mobility restriction). It should not be generalised to all NPIs. Risk communication and community engagement, safe and dignified burials, infection prevention and control in health facilities, isolation of contacts, and school closures are not modelled and cannot be evaluated by this analysis.

Several epidemiologic drivers known to influence Ebola transmission are not explicitly modelled: healthcare-associated transmission, funeral-associated transmission, heterogenous household transmission, delayed care-seeking, insecurity, community acceptance, and healthsystem disruption. Homogeneous mixing within districts likely overestimates transmission in low-density areas and underestimates it in high-density settlements. The DTM data cover only a 10-day window and eight border crossing points; movements through unofficial crossings or after the outbreak escalated may not be captured. Importations from neighbouring countries other than DRC were not considered. The model’s short-term validation is limited by the small number of observed data points. The small outbreak size produces low output variance, making precise quantitative sensitivity ranking unstable; we therefore interpret the sensitivity results qualitatively and rely on the total-order Sobol indices for the primary ranking. The extreme connectivity of affected zones—as quantified by Bangelesa et al., with Ituri Province having the second highest mobility intensity index nationally and the three epicentre health zones all exceeding the 95th percentile—makes contact tracing extraordinarily resource-intensive. The absence of a licensed vaccine against BDBV eliminates the ring-vaccination strategy that has been effective in all six preceding Ebola outbreaks in DRC caused by Zaire ebolavirus. Rapid strain-specific characterisation of biological parameters through contact tracing and serial interval analysis could yield greater returns than further refinement of operational protocols that are already performing adequately. Enhanced cross-border surveillance under International Health Regulations 2005 and the Africa CDC Public Health Emergency of Continental Security framework is crucial to harmonise detection and response across the DR Congo–Uganda– South Sudan tripoint. Preparedness planning may wish to evaluate pre-positioning response capacity at high-connectivity inland hubs while maintaining border surveillance at high-risk importation sites. These projections should not be interpreted as operational guidance without prospective validation. Dynamic mobility models using repeated DTM surveys or mobile phone data would capture how travel patterns evolve. Explicit modelling of household inaccessibility, nosocomial transmission, and community-level vaccination strategies would improve realism. Real-time updating of the model with new data would transform this retrospective analysis into a prospective decision-support tool.

## 5 Conclusions

The directed mobility network for Uganda is extremely unequal, with Kisoro serving as the primary gateway and conduit for EVD spread. Under the containment assumptions of this model, rapid implementation of a 7-1-7 response was associated with substantial containment to approximately 20 cases over 90 days, with additional modelled NPIs providing no statistically detectable reduction in case projections. These are scenario-conditional projections, not forecasts. They do not justify reducing border surveillance, nationwide preparedness, or community engagement; border districts remain the primary points of introduction, and NPIs outside those modelled here were not evaluated. Total-order Sobol indices identify contact-tracing coverage (ST = 0.69), latent period (ST = 0.35), and presymptomatic infectiousness (ST = 0.30) as the parameters with the largest effects on cumulative case counts. This framework provides a template for exploratory, network-based outbreak modelling applicable to other EVD strains, provided the results are interpreted within the corresponding assumptions.

## Data Availability

All the dataset supporting this analysis belong to the Ministry of Health, government of Uganda and access can be formally requested through the office of the Director General of Health Services.
The datasets and R code generated during and/or analysed during the current study are available from the corresponding author upon reasonable request. The DTM flow data can be officially requested from
IOM.

## List of Abbreviations

BDBV: Bundibugyo virus
CFR: Case-fatality ratio
CI: Confidence interval
DRC: Democratic Republic of the Congo
DTM: Displacement Tracking Matrix
EVD: Ebola virus disease
FMP: Flow Monitoring Point
HCW: Healthcare worker
IOM: International Organization for Migration
MoH: Ministry of Health
NPI: Non-pharmaceutical intervention
OFAT: One-factor-at-a-time
PHEIC: Public Health Emergency of International Concern
SEIR: Susceptible–Exposed–Infectious–Recovered
SI: Simulation interval
ST: Total-order Sobol index
UBOS: Uganda Bureau of Statistics
WHO: World Health Organization

## Declarations

### Ethics approval and consent to participate

Only anonymised, aggregated data were used; therefore ethical approval was not required.

### Consent for publication

Not applicable.

### Availability of data and materials

The datasets and R code generated during and/or analysed during the current study are available from the corresponding author upon reasonable request. The DTM flow data are publicly available from IOM.

### Authors’ contributions

AW, PM, BKN, SPS, MB, BM, LN, ML, TAS, BK, WS, WM, GT, BN, JK, MN, PAI, PK, ARA, AK, BL, BJK, MW, ANM, CO, and DA contributed to the study conception and design. AW, PM, BKN, and SPS contributed to data collection and analysis. AW and SPS contributed to model development and simulation. AW, PM, and BKN drafted the manuscript. All authors read and approved the final manuscript.

## Acknowledgements

We thank the Uganda Ministry of Health, the International Organization for Migration, and the Uganda Bureau of Statistics for data sharing that facilitated this analysis.

## Figure Legends

**Fig 6.** Morris screening: parameter importance ranked by µ ^∗^ (mean absolute elementary effect).

**Fig 7.** Total-order Sobol indices (Jansen estimator) with 95% bootstrap confidence intervals.

